# Safety and Exploratory Efficacy of Reduced β-Nicotinamide Mononucleotide Calcium Salt (NMNH-Ca) in Healthy Middle-Aged and Older Adults: A Randomized, Double-Blind, Placebo-Controlled Trial

**DOI:** 10.64898/2026.08.11.26360226

**Authors:** Jiayan Li, Ye Wang, Yanting Liang, Yun He, Eryong Jing, Qiang Shen, Jianjun Yu, Michael Chen, Chun Liang, Richard H. Kaszynski

## Abstract

Reduced nicotinamide mononucleotide (NMNH) is a reduced NAD⁺ precursor with reported NAD⁺-augmenting activity in preclinical models; however, controlled human data remain limited. This was a randomized, double-blind, placebo-controlled, parallel-group phase I trial evaluating oral NMNH-Ca in healthy adults aged 40-65 years. Eighty participants received placebo or NMNH-Ca 125, 250, or 500 mg once daily for 90 days. The primary objective was safety and tolerability. Whole-blood NAD⁺ was assessed as the key pharmacodynamic endpoint, including a 24-hour post-dose substudy, with biomarker-derived blood phenotypic age, treadmill-based six-minute walk distance, body mass index, and SF-36 domains analyzed as exploratory outcomes. NMNH-Ca was well tolerated at all doses, with no serious adverse events, treatment-related adverse events, or discontinuations. In the acute substudy, whole-blood NAD⁺ increased after single-dose NMNH-Ca, with peak mean concentrations at 12 hours. Over 90 days, NAD⁺ increased in a dose-related pattern; Day 90 mean changes from baseline were 2.33 ± 18.53 μM with placebo and 8.22 ± 10.25, 15.85 ± 11.16, and 39.90 ± 14.11 μM with NMNH-Ca 125, 250, and 500 mg, respectively. Exploratory analyses showed hypothesis-generating favorable signals in blood phenotypic age, treadmill-based six-minute walk distance, and health-related quality of life, most consistently at 500 mg. Oral NMNH-Ca was safe and pharmacodynamically active over 90 days, supporting larger and longer confirmatory trials with prespecified geroscience endpoints and tissue-relevant NAD⁺ metabolomics.

## 1. Introduction

Aging is the dominant biological substrate underlying multimorbidity, functional decline and late-life health system burdens (Chaudhary et al., 2023). At the cellular level, aging is not a single lesion but an interacting network of oxidative stress, mitochondrial dysfunction, inflammatory signaling and metabolic decline (Yang et al., 2024). These mechanisms are increasingly viewed as modifiable drivers of chronic disease rather than inevitable consequences of chronological time, resulting in the motivation of geroscientific interventions that target upstream biology across various organ systems (Kennedy et al., 2014).

Nicotinamide adenine dinucleotide (NAD^+^) occupies a central position within this framework. NAD^+^ is required for redox metabolism, mitochondrial function, DNA repair, sirtuin activity and immune-metabolic regulation (Yusri, Jose, Vermeulen, Tan, & Sorrentino, 2025). Age-related NAD^+^ decline and increased NAD^+^ consumption may impair mitochondrial resilience and stress response, providing a rationale for precursor-based NAD^+^ restoration (Poljšak, Kovač, & Milisav, 2022). Clinical studies of conventional NAD^+^ precursors, such as nicotinamide riboside (NR), nicotinamide mononucleotide (NMN), nicotinamide (NAM), and nicotinic acid (NA) have shown that oral supplementation can increase circulating or cellular NAD^+^ or related metabolites, although dose efficiency, tissue specificity and functional translation remain incompletely defined (Huang, 2022).

Reduced nicotinamide mononucleotide (NMNH), the reduced form of NMN, has emerged as a distinct NAD^+^ precursor candidate. Preclinical studies suggest that NMNH can increase intracellular NAD^+^ more rapidly and potently than oxidized precursor NMN (Y. Liu et al., 2021). Mechanistically, NMNH appears to enter NAD^+^ metabolism through pathways that are at least partly distinct from the canonical NRK and NAMPT-dependent routes used by conventional oxidized precursors. NMNH may be converted by nicotinamide mononucleotide adenylyltransferases (NMNAT) into NADH, which can subsequently be oxidized to NAD^+^, and may also undergo oxidation to NMN before conversion to NAD^+^ (Zapata-Pérez et al., 2021; Y. Liu et al., 2021). However, these mechanisms have been characterized primarily in preclinical systems, and their relevance after oral administration in humans remains to be established.

Given the limited human evidence, the present phase 1 randomized, double-blind, placebo-controlled trial evaluated oral NMNH-Ca supplementation in healthy adults aged 40-65 years. The primary objective was to assess safety and tolerability over 90 days. Whole-blood NAD^+^ concentration was evaluated as the key pharmacodynamic endpoint, with exploratory assessment of blood phenotypic age, body mass index (BMI), treadmill-based 6-minute walk distance and Short Form-36 (SF-36) health-related quality of life. Blood phenotypic age integrates routine clinical biomarkers associated with morbidity and mortality risk (Z. Liu et al., 2018), whereas the 6-minute walk test and SF-36 provide pragmatic functional and patient-reported outcomes with established clinical interpretability (Perera, Mody, Woodman, & Studenski, 2006) (Myhre et al., 2024). The present trial was designed to provide an initial clinical characterization of NMNH-Ca supplementation and to inform the design of larger studies evaluating whether NAD^+^ augmentation by reduced NAD^+^ precursors translates into reproducible geroscience-related effects.

## 2. Materials and Methods

### 2.1 Ethical Approval and Participants

The present randomized, double-blind, placebo-controlled, parallel-group clinical trial was reviewed and approved by the Independent Ethics Committees (IECs)/Institutional Review Boards (IRBs) of all participating sites, including Medstar Specialty Hospital Ethics Committee (Approval No. MEC/EP_RNM_001_24/14 APR 24, Bengaluru, Karnataka, India) and the Institutional Ethics Committee of Vinayaka Mission’s Medical College and Hospital (Approval No. VMMCH/IEC/2024/JULY/01, Karaikal, Puducherry, India). All study-related documents, including the clinical trial protocol, informed consent form (ICF), and case report forms (CRFs), were approved by the respective IECs/IRBs prior to study initiation. Study procedures were initiated only after obtaining all required ethical approvals. The trial was prospectively registered with ClinicalTrials.gov (Identifier: NCT06889740) and the Clinical Trials Registry of India (CTRI No.: CTRI/2024/06/068327) prior to participant enrollment. The study was conducted in accordance with the ethical principles of the Declaration of Helsinki (2013 revision), the International Council for Harmonisation (ICH) Guideline for Good Clinical Practice (E6 R2), and all applicable national regulatory requirements in India, including the New Drugs and Clinical Trials Rules (2019), the Indian Council of Medical Research (ICMR) National Ethical Guidelines for Biomedical and Health Research Involving Human Participants (2017), and the Good Clinical Practice Guidelines issued by the Central Drugs Standard Control Organization (CDSCO). The study adhered to CONSORT guidelines.

A total of 80 healthy adults aged 40-65 years who met the prespecified eligibility criteria were enrolled. Although this phase I trial was primarily designed to evaluate safety and tolerability, the target size was informed by a repeated-measures power calculation for longitudinal pharmacodynamic and exploratory endpoints. Using G*Power 3.1, a repeated-measures design with four groups and three measurement time points, assuming a moderate effect size (f = 0.25), two-sided α = 0.05, power (1 − β) = 0.9, correlation among repeated measures = 0.5 and nonsphericity correction ε = 0.75, indicated that 64 participants would be required. Allowing for an anticipated dropout rate of 20%, the enrollment target was set at 80 participants.

Eligible participants had a body mass index (BMI) between 18.5 and 35.0 kg/m²who maintained relatively stable diet and lifestyle habits. No monetary compensation is provided in the study. Individuals were excluded if they were using NAD⁺-related supplements (e.g., NMN, NR, NA) or statin medications, had used tobacco or recreational drugs within the previous six months, or had abnormal laboratory findings or medical conditions that could interfere with the study participation as judged by the investigator. Additional exclusion criteria included a history of atherosclerotic, cardiopulmonary, or significant psychiatric disorders, substance abuse, or hypersensitivity to study product components. Participants were also excluded if they were unwilling to discontinue vitamin or dietary supplements prior to study initiation, planned to undertake weight loss interventions, or had lifestyle constraints incompatible with protocol adherence. Women who were pregnant, breastfeeding, or had a positive pregnancy test were not eligible. Individuals with recent participation in another clinical study, inability to provide venous blood samples, or a diagnosis of cancer within the past five years were also excluded.

### 2.2 Study Design

The investigational product was a food-grade capsule containing NMNH-Ca, marketed as UthPeak® NMNH-Ca. NMNH calcium salt exhibits greater stability and lower hygroscopicity than both free NMNH and NMNH disodium salt, and its simpler preparation process supports large-scale production. The product was developed and manufactured by Effepharm (Shanghai) Co. Ltd. under Good Manufacturing Practice and Good Clinical Practice standards. The investigational product was subsequently transferred to IDD Research Solutions Pvt. Ltd., where secondary packaging, labelling and clinical trial supply management was performed in accordance with applicable regulatory requirements and GCP standards. Matching placebo capsules were identical in appearance, packaging and labeling to the active investigational product.

As shown in Figure 1, eligible participants were randomly assigned to one of four parallel groups: placebo, NMNH-Ca 125 mg, NMNH-Ca 250 mg, and NMNH-Ca 500 mg, with a planned allocation ratio of 1:1:1:1. The final analyzed group sizes were 21, 18, 20, and 21 participants, respectively. The overall study design, treatment allocation, dosing schedule, and assessment timeline are summarized in Figure 1. Both participants and investigators remained blinded to group allocation throughout the study. Treatment assignments were identified using coded labels, and the randomization codes were securely maintained and not disclosed until completion of the study and database review.

**Figure 1.**
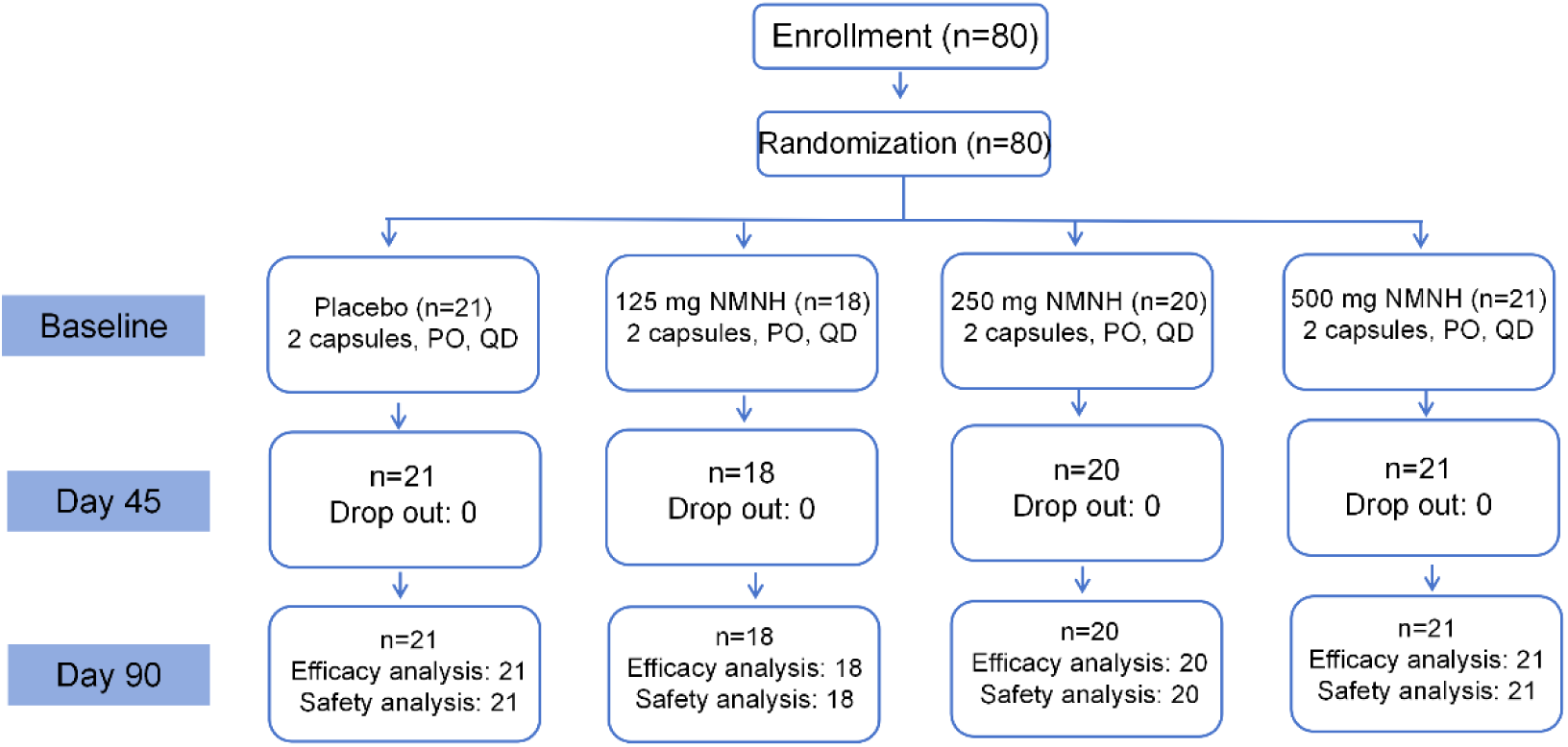
Flow diagram of the participants. PO = orally, QD = once daily dosing. Flow diagram showing participant enrollment, randomization, treatment allocation, dosing schedule, follow-up visits, and analysis populations. Participants were randomized to placebo or NMNH-Ca 125 mg, 250 mg, or 500 mg once daily for 90 days. PO, oral administration; QD, once daily.

Participants received the assigned investigational product or placebo once daily in the morning, immediately after breakfast, for 90 consecutive days. Participants were instructed to maintain their usual diet, physical activity and lifestyle habits throughout the intervention period. Compliance was monitored through scheduled follow-up and participant reporting. Study assessments were performed at baseline, Day 45, and Day 90.

The primary objective of this phase I trial was to evaluate the safety and tolerability of repeated oral NMNH-Ca supplementation. Whole-blood NAD⁺ concentration was assessed as the key pharmacodynamic endpoint, including both an acute 24-hour post-dose assessment and longitudinal measurements over 90 days. Exploratory geroscience-related endpoints included blood phenotypic age, six-minute walk test distance, body mass index, and SF-36 health-related quality-of-life scores.

### 2.3 Determination of NAD⁺ Concentration in Whole-blood

Whole-blood NAD⁺ concentrations were measured using a commercially available NAD^+^ microplate assay according to the manufacturer’s instructions (MyBioSource, USA; Catalog #MBS822347). Acute pharmacodynamic responses were assessment in a subset of 40 participants, consisting of 10 participants from each treatment group who consented to intensive sampling. On Day 1, blood samples were collected before dosing and at 0.5, 1, 2, 4, 6, 12, and 24 hours after dosing. Longitudinal NAD⁺ concentrations were assessed in all participants at baseline, Day 45, and Day 90.

### 2.4 Estimated Blood Phenotypic Age Assessment

Estimated blood phenotypic age was measured using a biomarker based algorithm incorporating the following: albumin (g/dL), creatinine (mg/dL), glucose (mg/dL), C-reactive protein (CRP, mg/L), lymphocytes percentage (%), mean corpuscular volume (MCV, fL), red blood cell distribution width (RDW, %), alkaline phosphatase (U/L), white blood cells (WBC, ×10³/uL) and chronological age. Measurements were conducted at baseline, Day 45, and Day 90 and phenotypic age was calculated using a spreadsheet-based implementation of the published algorithm (Levine et al., 2018).

Because estimated blood phenotypic age is a biomarker-derived estimate rather than a direct measure of biological aging, this endpoint was analyzed as exploratory. A post hoc responder analyses was performed by which an estimated blood phenotypic age responder was defined as a participant achieving a reduction of at least 3 years in estimated blood phenotypic age from baseline to Day 90. This threshold was selected to exceed expected short-term biological and analytical variability and to provide a conservative and clinically interpretable benchmark at the individual level.

### 2.5 Six-Minute Walking Test (6MWT)

The 6MWT was used as an exploratory assessment of submaximal functional capacity and walking endurance. Testing was conducted at baseline, Day 45, and Day 90 using a motorized treadmill, and total distance covered over six minutes was recorded in meters using the treadmill’s digital odometer.

A post hoc responder analyses was performed using a threshold of at least 50 m improvement from baseline to Day 90. This threshold was selected as it exceeds commonly reported minimal clinically important difference of approximately 25 to 30 m and was intended to identify robust functional improvement rather than minor measurement variation (Perera et al., 2006) (Myhre et al., 2024).

### 2.6 Health-Related Quality of Life (HRQOL)

HRQOL was evaluated at Day 0, 45 and 90 using the 36-Item Short Form Health Survey (SF-36) (McHorney, Ware Johne, & ANASTASIAE, 1993). The eight domains including physical functioning, bodily pain, role limitations due to physical health problems, role limitations due to personal or emotional problems, emotional well-being, social functioning, energy/fatigue, and general health perceptions. Scores range from 0 to 100, with higher scores indicating a more favorable health state.

### 2.7 Statistical Analysis

Statistical analyses were performed using IBM SPSS Software, version 29.0.2.0. Two-sided p-value < 0.05 were considered statistically significant. Because this phase I trial was primarily designed to evaluate safety and tolerability, analysis of pharmacodynamic and exploratory endpoints were considered supportive and hypothesis-generating.

Safety outcomes were summarized by treatment group, including adverse events, serious adverse events, treatment-related adverse events, discontinuations, severity, outcome, and investigator-assessed causality. Baseline characteristics were summarized descriptively. Between-group baseline comparisons were performed using unpaired t-tests for continuous variables and chi-square or Fisher’s exact tests for categorical variables as appropriate.

For longitudinal outcomes, including blood NAD⁺ concentrations, 6MWT distance, estimated blood phenotypic age and SF-36 scores, changes from baseline were calculated at each post-baseline visit. Within-group changes were assessed using paired t-tests. Between-group differences over time between each NMNH-Ca dose group and placebo were assessed using a mixed model for repeated measures (MMRM), with treatment group, visit, and treatment-by-visit interaction as fixed effects. Multiplicity across comparisons of each NMNH-Ca dose group versus placebo was controlled using Dunnett’s test.

Post hoc placebo-adjusted Day 90 treatment effects were estimated for each endpoint. Standardized between-group effect sizes were calculated as Hedges’ g using the placebo-adjusted mean change divided by the pooled standard deviation with small-sample bias correction. Corresponding 95% confidence intervals were estimated and *p* values were adjusted using the Holm method for dose-group comparisons within each outcome.

## 3. Results

### 3.1 Participant Disposition and Baseline Characteristics

A total of 80 eligible participants were randomized to placebo, NMNH-Ca 125 mg, NMNH-Ca 250 mg and NMNH-Ca 500 mg, with final group sizes of 21, 18, 20 and 21 participants, respectively. All participants completed the 90-day intervention and were included in the analysis. Participants were 40 to 65 years of age, 69/80 were male and all were Asian. Baseline demographic and clinical characteristics, including age, sex distribution, and anthropometric measures, were generally comparable across treatment groups (Table 1). Most participants reported mild or moderate physical activity and no participants reported current tobacco or alcohol use.

### 3.2 Acute Whole-Blood NAD^+^ Pharmacodynamic Response After Single-Dose Administration

A subset of 40 participants, with 10 participants per group, was included in the 24-hour NAD^+^ pharmacodynamic assessment. Following a single oral dose of NMNH-Ca, whole-blood NAD^+^ concentrations increased after administration and reached peak mean concentrations at 12 hours, with values of 49.0 μM, 50.9 μM, and 67.1 μM in the 125 mg, 250 mg, and 500 mg groups, respectively. By 24 hours, NAD^+^ concentrations had declined but remained higher than those observed in the placebo group, which showed no apparent post-dose increase (Figure 2A). Baseline-corrected 24-hour NAD^+^ pharmacodynamic response was higher in NMNH-Ca-treated participants than in placebo-treated participants, with the greatest exposure observed in the 500 mg group (Figure 2B). These findings suggest that once-daily dosing is sufficient to maintain elevated NAD^+^ levels over a 24-hour period, supporting the dosing regimen used in this study.

**Figure 2.**
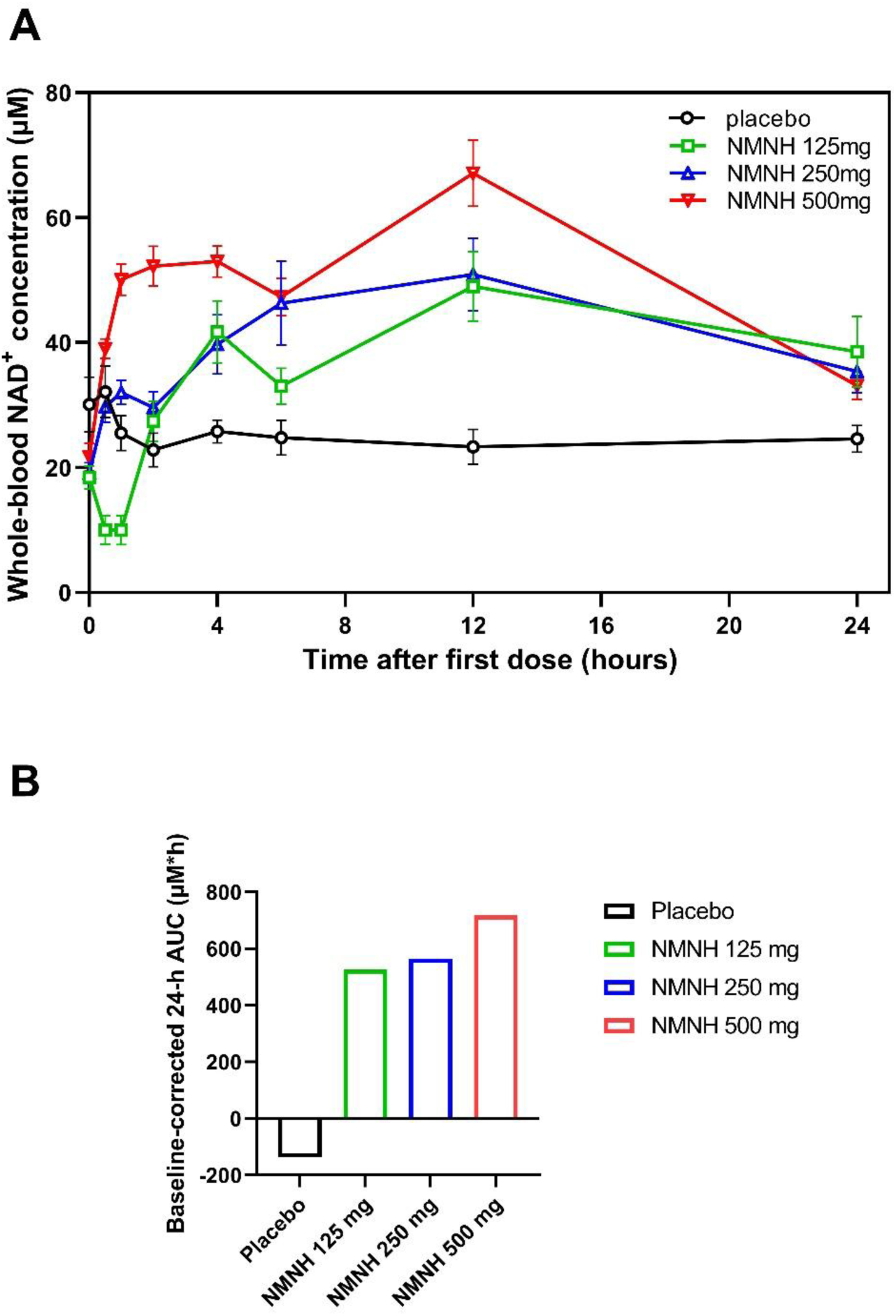
Acute whole-blood NAD+ pharmacodynamic response after single-dose NMNH-Ca administration. (**A**) Whole-blood NAD+ concentrations were measured before dosing and at 0.5, 1, 2, 4, 6, 12, and 24 hours after a single oral dose of placebo or NMNH-Ca at 125 mg, 250 mg, or 500 mg. Data are presented as mean ± SEM. (**B**) Baseline-corrected incremental AUC_0-24 h_ for whole-blood NAD^+^ concentration after single-dose administration. Incremental AUC_0-24 h_ was calculated after subtraction of each participant’s predose NAD^+^ value. Bars represent group means.

**Table 1.** Baseline demographic and clinical characteristics by treatment group.

| Characteristic | Placebo | NMNH-Ca<br>125 mg | NMNH-Ca<br>250 mg | NMNH-Ca<br>500 mg | Overall |
| --- | --- | --- | --- | --- | --- |
| Participants, n | 21 | 18 | 20 | 21 | 80 |
| Age, years, mean (SD) | 46.52 (5.15) | 47.06 (5.46) | 47.20 (7.13) | 46.43 (5.11) | 46.79 (5.67) |
| Female sex, n (%) | 1 (4.8) | 2 (11.1) | 3 (15.0) | 5 (23.8) | 11 (13.8) |
| Male sex, n (%) | 20 (95.2) | 16 (88.9) | 17 (85.0) | 16 (76.2) | 69 (86.3) |
| Asian race, n (%) | 21 (100.0) | 18 (100.0) | 20 (100.0) | 21 (100.0) | 80 (100.0) |
| BMI, kg/m<sup>2</sup>, mean (SD) | 24.40 (2.27) | 24.73 (1.71) | 24.92 (2.07) | 25.00 (2.29) | 24.76 (2.08) |
| Height, m, mean (SD) | 1.68 (0.18) | 1.68 (0.07) | 1.69 (0.08) | 1.65 (0.09) | 1.68 (0.11) |
| Weight, kg, mean (SD) | 72.00 (5.48) | 70.13 (5.06) | 71.23 (4.99) | 68.04 (5.11) | 70.35 (5.30) |
| No physical activity, n (%) | 0 (0.0%) | 0 (0.0%) | 1 (5.0%) | 1 (4.8%) | 2 (2.5%) |
| Mild physical activity, n (%) | 14 (66.7%) | 13 (72.2%) | 6 (30.0%) | 12 (57.1%) | 45 (56.3%) |
| Moderate physical activity, n (%) | 7 (33.3%) | 5 (27.8%) | 13 (65.0%) | 8 (38.1%) | 33 (41.3%) |
| Never smoked, n (%) | 21 (100.0) | 18 (100.0) | 20 (100.0) | 21 (100.0) | 80 (100.0) |
| Alcohol never-use, n (%) | 21 (100.0) | 18 (100.0) | 20 (100.0) | 21 (100.0) | 80 (100.0) |

Values are presented as mean ± SD for continuous variables and n (%) for categorical variables. BMI, body mass index; NMNH-Ca, reduced $\beta$ -nicotinamide mononucleotide calcium salt. Percentages may not total 100 because of rounding.

### 3.3 Longitudinal Whole-Blood NAD^+^ Response During 90-Day Supplementation

Whole-blood NAD⁺ concentrations were assessed at baseline, Day 45, and Day 90. Baseline NAD^+^ concentrations were broadly comparable across treatment groups (Table 2). During the 90-day intervention period, NAD^+^ concentrations remained relatively stable in the placebo group, whereas NMNH-Ca supplementation was associated with time-and dose-related increases (Figure 3A). At Day 90, mean changes from baseline were 2.33 ± 18.53 μM in the placebo group, compared with 8.22 ± 10.25 μM, 15.85 ± 11.16 μM, and 39.90 ± 14.11 μM in the 125 mg, 250 mg, and 500 mg groups, respectively. The 250 mg and 500 mg groups showed significant placebo-adjusted increases at Day 90 (p < 0.001), with the largest effect observed in the 500 mg group (Figure 3B). In the 500 mg group, mean NAD^+^ concentration increased from 19.43 ± 6.54 μM at baseline to 59.33 ± 13.27 μM at Day 90.

**Figure 3.**
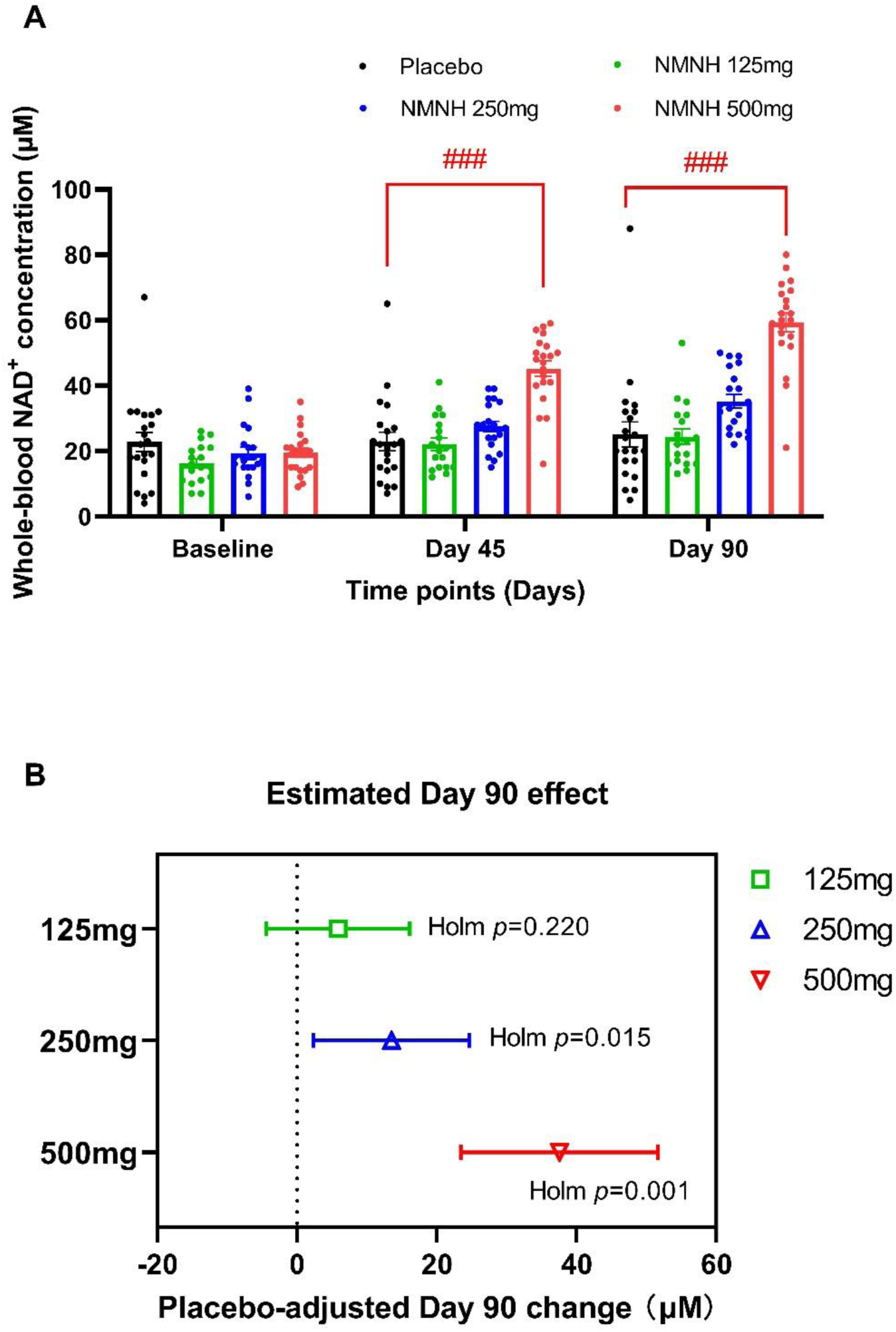
Effects of NMNH-Ca supplementation on whole-blood NAD^+^ concentrations. (**A**) The whole-blood NAD^+^ concentrations are shown at baseline, Day 45 and Day 90 for the placebo, NMNH-Ca 125 mg, 250 mg, and 500 mg groups. Data are presented as mean ± SEM. ###*p* < 0.001 versus placebo. (**B**) Placebo-adjusted Day 90 changes from baseline in whole-blood NAD^+^ concentration for each NMNH-Ca dose group. Points indicate estimated treatment effects, and horizontal bars indicate 95% CIs. The *p* values were adjusted using the Holm method for dose-group comparisons within the endpoint.

**Table 2.**
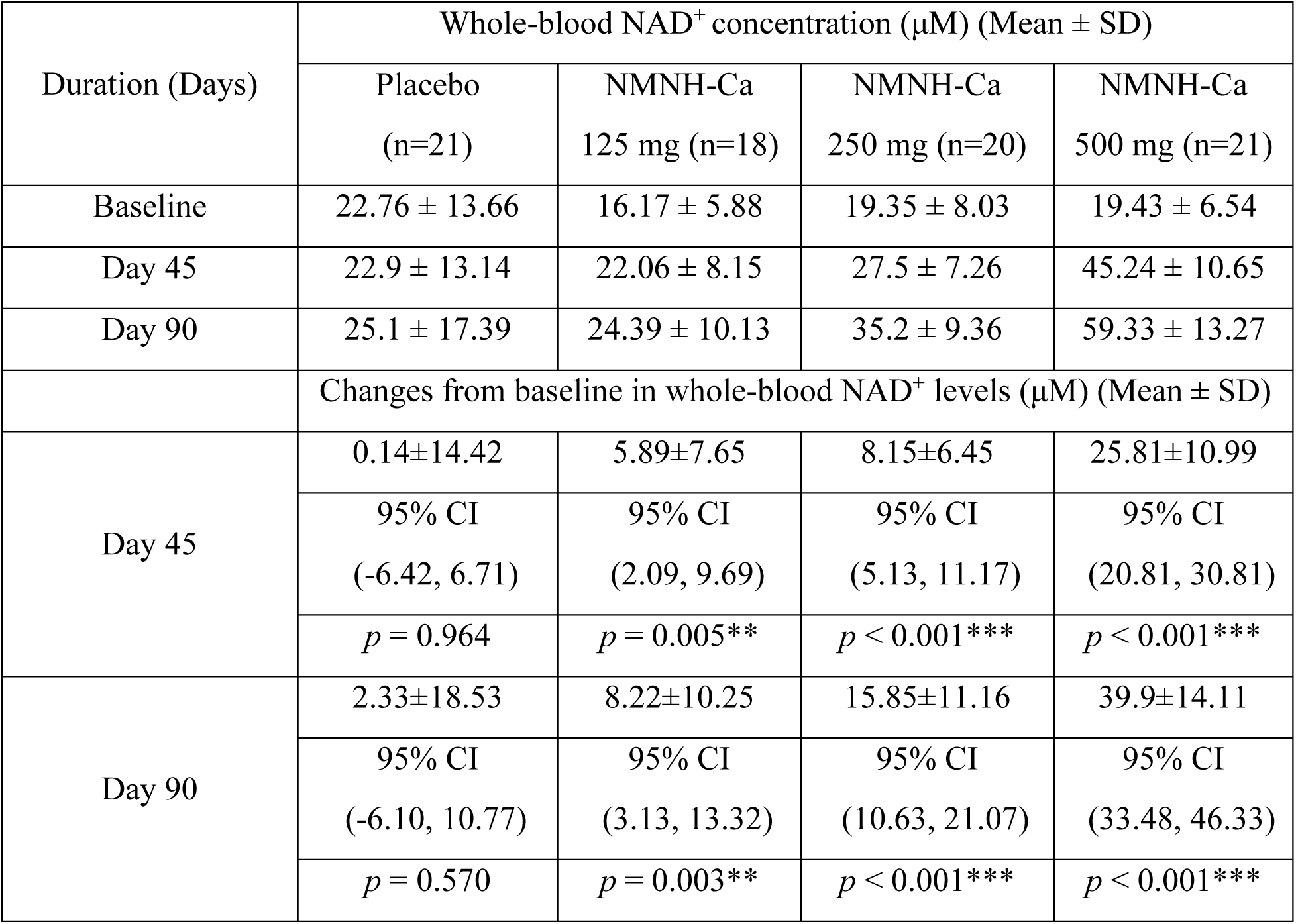
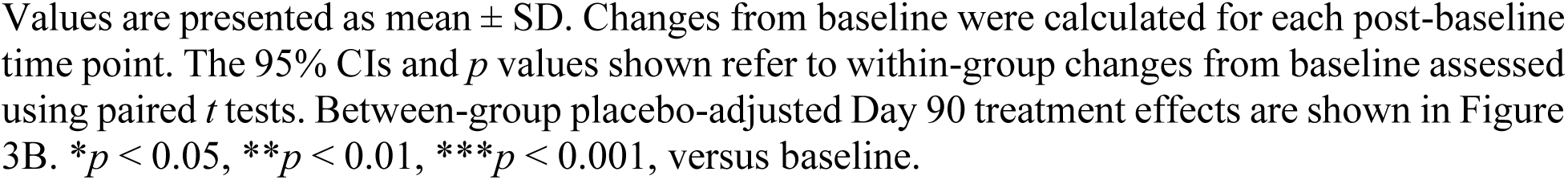
Whole-blood NAD⁺ concentrations and changes from baseline during 90-day supplementation.

| Duration (Days) | Whole-blood NAD<sup>+</sup> concentration (μM) (Mean ± SD) | | | |
| --- | --- | --- | --- | --- |
|  | Placebo<br>(n=21) | NMNH-Ca<br>125 mg (n=18) | NMNH-Ca<br>250 mg (n=20) | NMNH-Ca<br>500 mg (n=21) |
| Baseline | 22.76 ± 13.66 | 16.17 ± 5.88 | 19.35 ± 8.03 | 19.43 ± 6.54 |
| Day 45 | 22.9 ± 13.14 | 22.06 ± 8.15 | 27.5 ± 7.26 | 45.24 ± 10.65 |
| Day 90 | 25.1 ± 17.39 | 24.39 ± 10.13 | 35.2 ± 9.36 | 59.33 ± 13.27 |
| Changes from baseline in whole-blood NAD<sup>+</sup> levels (μM) (Mean ± SD) | | | | |
| Day 45 | 0.14±14.42 | 5.89±7.65 | 8.15±6.45 | 25.81±10.99 |
|  | 95% CI<br>(-6.42, 6.71) | 95% CI<br>(2.09, 9.69) | 95% CI<br>(5.13, 11.17) | 95% CI<br>(20.81, 30.81) |
| | $p = 0.964$ | $p = 0.005^{**}$ | $p < 0.001^{***}$ | $p < 0.001^{***}$ |
| Day 90 | 2.33±18.53 | 8.22±10.25 | 15.85±11.16 | 39.9±14.11 |
|  | 95% CI<br>(-6.10, 10.77) | 95% CI<br>(3.13, 13.32) | 95% CI<br>(10.63, 21.07) | 95% CI<br>(33.48, 46.33) |
| | $p = 0.570$ | $p = 0.003^{**}$ | $p < 0.001^{***}$ | $p < 0.001^{***}$ |

Values are presented as mean ± SD. Changes from baseline were calculated for each post-baseline time point. The 95% CIs and $p$ values shown refer to within-group changes from baseline assessed using paired $t$ tests. Between-group placebo-adjusted Day 90 treatment effects are shown in Figure 3B. \* $p < 0.05$ , \*\* $p < 0.01$ , \*\*\* $p < 0.001$ , versus baseline.

### 3.4 Exploratory Changes in Estimated Blood Phenotypic Age

Estimated blood phenotypic age was assessed at baseline, Day 45, and Day 90 (Table 3 and Figure 4A). At Day 90, estimated blood phenotypic age increased by 2.62 ± 4.03 years in the placebo group, whereas changes in the 125 mg, 250 mg, and 500 mg groups were 0.02 ± 5.58 years, −0.85 ± 10.15 years, and −5.18 ± 3.22 years, respectively. The most pronounced change was observed in the 500 mg group, in which mean estimated blood phenotypic age decreased from 45.24 ±7.99 years at baseline to 40.06 ±5.89 years at Day 90.

**Figure 4.**
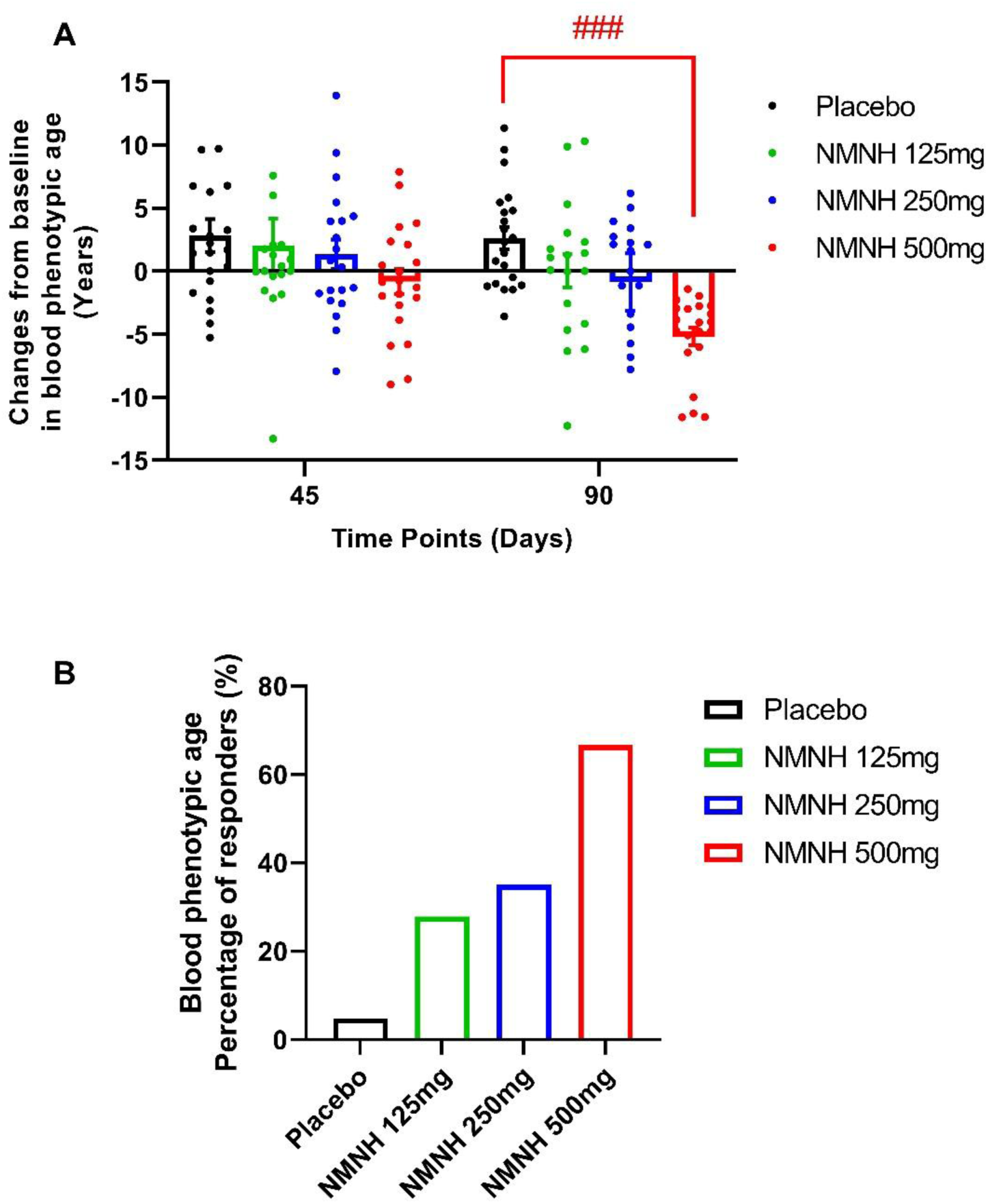
Exploratory changes in estimated blood phenotypic age during 90-day NMNH-Ca supplementation. (**A**) Changes from baseline in estimated blood phenotypic age were assessed at Day 45 and Day 90 in participants receiving placebo or NMNH-Ca at doses of 125 mg, 250 mg, or 500 mg. Data are shown as group means with SEM. Lower values indicate a reduction in biomarker-derived phenotypic age. ###*p* < 0.001 versus placebo. (**B**) Responder analysis showing the proportion of participants achieving a reduction of at least 3 years in blood phenotypic age from baseline to Day 90.

**Table 3.** Blood phenotypic age and changes from baseline during 90-day supplementation.

| Duration (Days) | Blood phenotypic age (years) (Mean ± SD) | | | |
| --- | --- | --- | --- | --- |
|  | Placebo<br>(n = 21) | NMNH-Ca<br>125 mg<br>(n = 18) | NMNH-Ca<br>250mg<br>(n = 20) | NMNH-Ca<br>500 mg<br>(n = 21) |
| Baseline | 46.68 ± 5.47 | 45.22 ± 6.72 | 45.64 ± 7.09 | 45.24 ± 7.99 |
| Day 45 | 49.52 ± 7.86 | 47.26 ± 7.49 | 46.99 ± 7.48 | 44.43 ± 5.98 |
| Day 90 | 49.31 ± 5.48 | 45.23 ± 7.10 | 44.79 ± 8.67 | 40.06 ± 5.89 |
| Changes from baseline in blood phenotypic age (years) (Mean ± SD) | | | | |
| Day 45 | 2.84 ± 5.92 | 2.04 ± 9.02 | 1.35 ± 5.19 | -0.81 ± 4.43 |
|  | 95% CI<br>(0.14, 5.53) | 95% CI<br>(-2.44, 6.53) | 95% CI<br>(-1.08, 3.78) | 95% CI<br>(-2.83, 1.20) |
| | $p = 0.040^*$ | $p = 0.350$ | $p = 0.260$ | $p = 0.411$ |
| Day 90 | 2.62 ± 4.03 | 0.02 ± 5.58 | -0.85 ± 10.15 | -5.18 ± 3.22 |
|  | 95% CI | 95% CI | 95% CI | 95% CI |
|  | (0.79, 4.46) | (-2.76, 2.79) | (-5.60, 3.90) | (-6.64, -3.71) |
| | $p = 0.007^{***}$ | $p = 0.991$ | $p = 0.712$ | $p < 0.001^{***}$ |

Values are presented as mean ± SD. Changes from baseline were calculated for each post-baseline time point. The 95% CIs and p values shown refer to within-group changes from baseline assessed using paired *t* tests. Lower values and negative changes indicate lower biomarker-derived phenotypic age. \* $p < 0.05$ and \*\*\* $p < 0.001$ versus baseline.

A responder analysis, defined as a reduction of at least 3 years in estimated blood phenotypic age from baseline to Day 90, showed a graded pattern across treatment groups. Responder rates were 4.8% in the placebo group, 27.8% in the 125 mg group, 35.0% in the 250 mg group, and 66.7% in the 500 mg group (Figure 4B). These findings suggest a dose-related pattern in this exploratory biomarker-derived endpoint.

### 3.5 Exploratory Functional and Health-Related Quality-of-Life Outcomes

Physical function was assessed using the 6MWT. Baseline walking distance was similar across treatment groups (Table 4). At Day 90, mean walking distance increased by approximately 25 m in the placebo group, compared with approximately 12 m, 45 m, and 114 m in the 125 mg, 250 mg, and 500 mg groups, respectively. The largest improvement was observed in the 500 mg group, where mean distance increased from 813 ± 211 m at baseline to 927 ± 193 m at Day 90 (Figure 5A).

**Figure 5.**
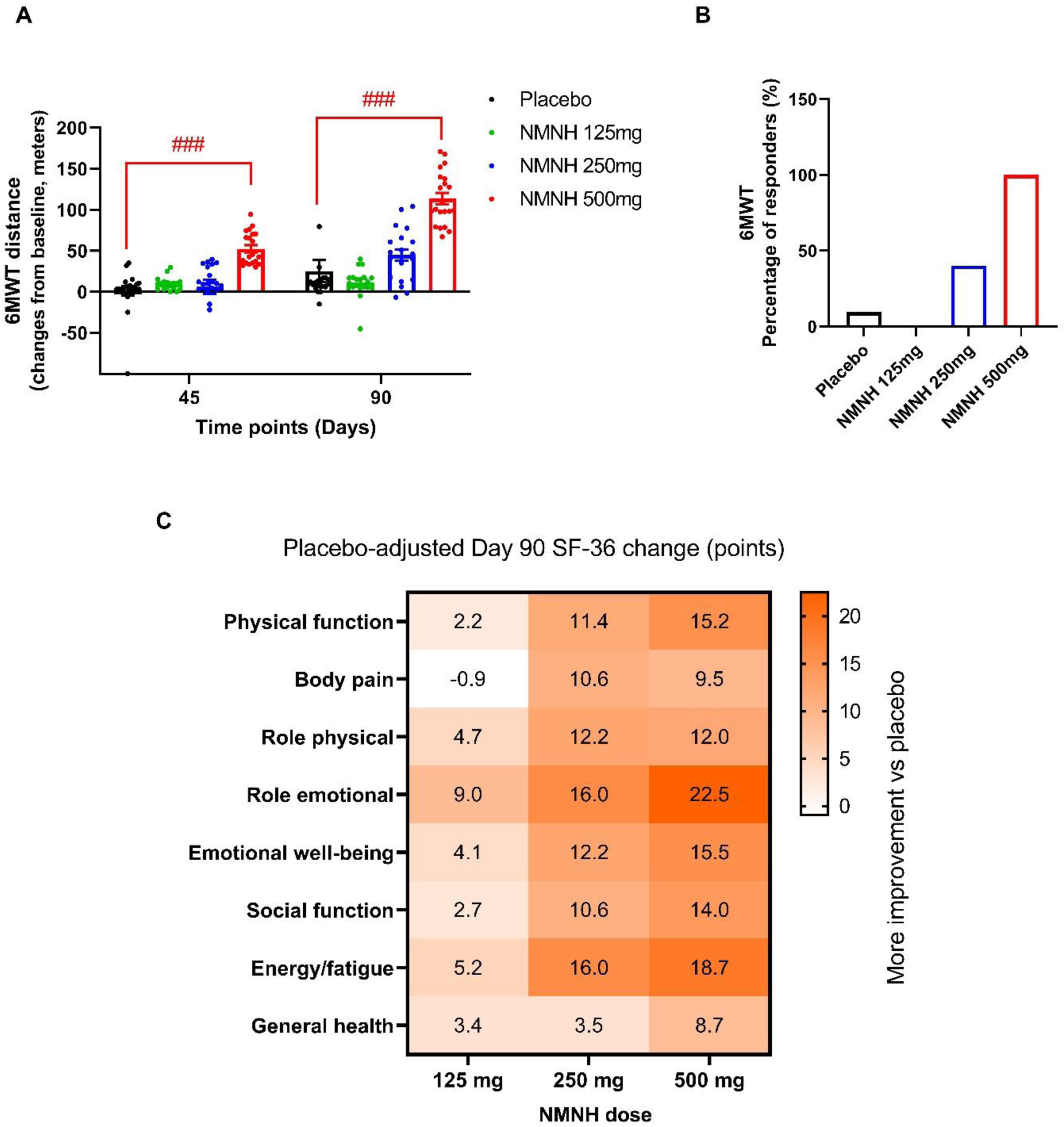
Exploratory functional and health-related quality-of-life outcomes during 90-day NMNH-Ca supplementation. (**A**) Change from baseline in six-minute walk test distance at Day 45 and Day 90 in participants receiving placebo or NMNH-Ca at 125 mg, 250 mg, or 500 mg once daily. Data are presented as mean ± SEM. ###*p* < 0.001 versus placebo. (**B**) Responder analysis showing the proportion of participants achieving an improvement of at least 50 m in six-minute walk test distance from baseline to Day 90. (C) Heatmap of placebo-adjusted Day 90 changes in SF-36 domain scores across NMNH-Ca dose groups. Values within cells represent estimated treatment differences versus placebo. Higher SF-36 scores indicate more favorable health-related quality of life, and warmer colors indicate greater improvement relative to placebo.

**Table 4.** Treadmill-based 6MWT distance and changes from baseline during 90-day supplementation.

| Duration (Days) | Mean total distance covered (meters) (Mean ± SD) | | | |
| --- | --- | --- | --- | --- |
|  | Placebo<br>(n = 21) | NMNH-Ca<br>125 mg<br>(n = 18) | NMNH-Ca<br>250 mg<br>(n = 20) | NMNH-Ca<br>500 mg<br>(n = 21) |
| Baseline | 854 ± 212 | 851 ± 215 | 833 ± 213 | 813 ± 211 |
| Day 45 | 855 ± 218 | 861 ± 214 | 839 ± 216 | 866 ± 207 |
|  | 95% CI<br>(-10.66, 13.13) | 95% CI<br>(5.680, 13.54) | 95% CI<br>(-12.48, 24.78) | 95% CI<br>(43.66, 61.58) |
| | $p = 0.8303$ | $p = 0.0569$ | $p = 0.4980$ | $p < 0.0001^{***}$ |
| Day 90 | 879 ± 230 | 863 ± 220 | 878 ± 210 | 927 ± 193 |
|  | 95% CI<br>(-2.417, 53.08) | 95% CI<br>(2.652, 20.57) | 95% CI<br>(30.13, 59.87) | 95% CI<br>(99.41, 128.5) |
| | $p = 0.0714$ | $p = 0.0141^*$ | $p < 0.0001^{***}$ | $p < 0.0001^{***}$ |

Values are presented as mean ± SD. Changes from baseline were calculated for each post-baseline time point. The 95% CIs and $p$ values shown refer to within-group changes from baseline assessed using paired $t$ tests. $^*p < 0.05$ , $^{***}p < 0.001$ , versus baseline.

In the responder analysis, defined as an improvement of at least 50 m from baseline to Day 90, responder rates were 9.5% in the placebo group, 0.0% in the 125 mg group, 40.0% in the 250 mg group, and 100.0% in the 500 mg group (Figure 5B). These findings suggest a dose-related pattern in this exploratory functional endpoint.

Health-related quality of life was assessed using the SF-36. Domain-level data are provided in Supplementary Document 1. At Day 90, the 500 mg group showed improvements across all eight SF-36 domains, while the 250 mg group showed generally smaller improvements and the 125 mg group showed more variable changes (Figure 5C). These patient-reported outcomes should be interpreted as exploratory.

### 3.6 Integrated post hoc placebo-adjusted Day 90 treatment-effect estimates

An integrated post hoc analysis was performed to compare standardized placebo-adjusted Day 90 treatment effects across whole-blood NAD^+^ concentration, estimated blood phenotypic age, 6MWT, and BMI (Figure 6). For whole-blood NAD⁺, placebo-adjusted mean Day 90 differences were +5.89 μM, +13.52 μM, and +37.57 μM in the NMNH-Ca 125 mg, 250 mg, and 500 mg groups, respectively. The corresponding standardized treatment-effect estimates were Hedges’ g = 0.38, 0.86, and 2.24, with Holm-adjusted p values of 0.238, 0.015, and <0.001, respectively.

**Figure 6.**
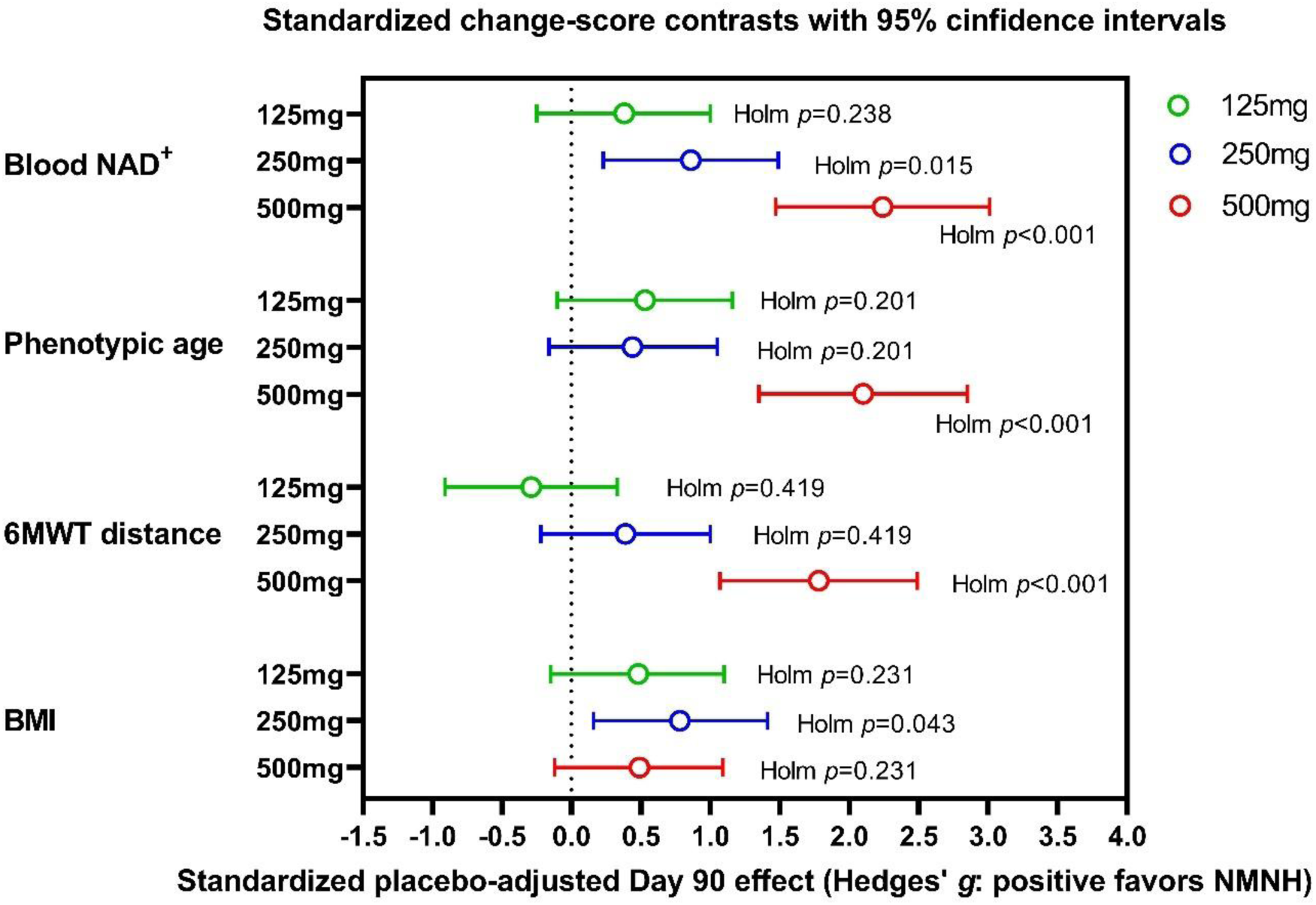
Integrated post hoc placebo-adjusted Day 90 standardized treatment-effect estimates. Standardized placebo-adjusted Day 90 treatment effects are shown for whole-blood NAD^+^ concentration, blood phenotypic age, 6MWT distance, and BMI in participants receiving NMNH-Ca at 125 mg, 250 mg, or 500 mg once daily. Points indicate Hedges’ g estimates, and horizontal bars indicate 95% CIs. Effect directions were oriented so that positive values indicate a favorable effect of NMNH-Ca relative to placebo for NAD^+^ concentration, blood phenotypic age, and six-minute walk distance. BMI is shown as a body-weight reference endpoint. Multiplicity across dose-group comparisons within each outcome was adjusted using the Holm method, with Holm-adjusted *p* values shown.

For estimated blood phenotypic age, placebo-adjusted Day 90 differences were −2.60 years, −3.47 years, and −7.80 years in the 125 mg, 250 mg, and 500 mg groups, respectively, where lower values indicate a more favorable biomarker-derived phenotypic-age profile. After orienting effect direction so that positive Hedges’ *g* values favor NMNH-Ca, the corresponding standardized effects were Hedges’ *g* = 0.53, 0.44, and 2.10, with Holm-adjusted p values of 0.201, 0.201, and <0.001, respectively.

Treadmill-based 6MWT distance placebo-adjusted Day 90 differences were −13.7 m, +19.7 m, and +88.6 m in the 125 mg, 250 mg, and 500 mg groups, respectively. The corresponding standardized treatment effects were Hedges’ *g* = −0.29, 0.39, and 1.79, with Holm-adjusted p values of 0.419, 0.419, and <0.001, respectively.

BMI was included as a body-weight reference endpoint. Standardized BMI contrasts were not dose-monotonic, with Hedges’ g values of approximately 0.48, 0.78, and 0.48 in the 125 mg, 250 mg, and 500 mg groups, respectively, and Holm-adjusted p values of 0.231, 0.043, and 0.231.

### 3.7 Safety and Tolerability

NMNH-Ca was well tolerated across all dose groups during the 90-day intervention period. Six mild treatment-emergent adverse events were reported: body pain in one participant each in the 125 mg and 250 mg groups, fever in one participant each in the 125 mg and 500 mg groups, and headache in one participant each in the placebo and 500 mg groups (Table 5). All events resolved without treatment, and none were considered related to NMNH-Ca supplementation. No serious adverse events, treatment-related adverse events, or discontinuations due to adverse events were reported.

**Table 5.**
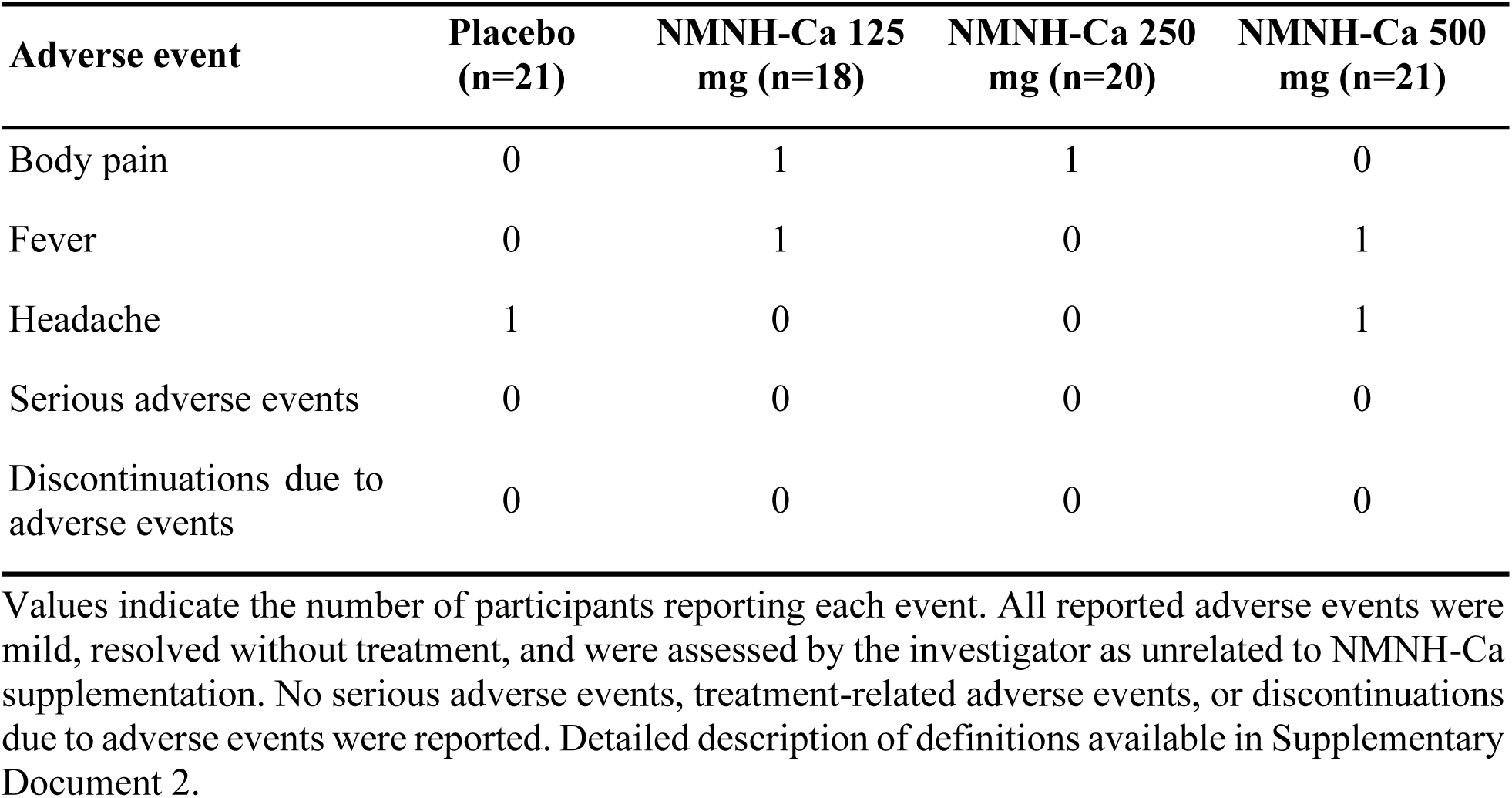
Treatment-emergent adverse events during the 90-day intervention period.

## 4. Discussion

This randomized, double-blind, placebo-controlled, parallel-group phase I trial provides initial clinical evidence that oral NMNH-Ca supplementation is generally well-tolerated and pharmacodynamically active in healthy adults aged 40-65 years. Across daily doses of NMNH-Ca 125, 250, or 500 mg administered for 90 days, NMNH-Ca was not associated with serious adverse events, treatment-related adverse events, or discontinuations. The study further demonstrated rapid and sustained increases in whole-blood NAD⁺ concentrations, with the largest response observed in the 500 mg group. Exploratory analyses suggested favorable dose-related patterns in blood phenotypic age, 6MWT, and health-related quality-of-life measures. These findings align with the geroscience framework, which views aging biology as a modifiable network of upstream mechanisms contributing to chronic disease vulnerability rather than as an inevitable consequence of chronological aging alone (López-Otín, Blasco, Partridge, Serrano, & Kroemer, 2013) (Kennedy et al., 2014).

NAD^+^ metabolism is a central target in aging biology because of its involvement in mitochondrial function, redox homeostasis, DNA repair, sirtuin activity, PARP signaling, and cellular stress responses (Yusri et al., 2025). Several NAD^+^ precursors, including nicotinamide riboside (Elhassan et al., 2019) and nicotinamide mononucleotide (Okabe et al., 2022), have been evaluated clinically and shown to increase circulating or tissue NAD^+^-related metabolites with generally favorable safety profiles. NMNH differs from these oxidized precursors by existing in a reduced chemical state and has been reported in preclinical studies to increase NAD^+^ more efficiently than NMN through partially distinct metabolic routes (Y. Liu et al., 2021) (Zapata-Pérez et al., 2021). In this context, the present study extends prior preclinical observations by showing that orally administered NMNH-Ca increases whole-blood NAD^+^ in humans after both single-dose administration and repeated supplementation.

The pharmacodynamic signal is biologically relevant in the context of NAD^+^ biology. In the 24-hour substudy, NMNH-Ca increased whole-blood NAD^+^ after a single oral dose, with peak concentrations observed at 12 hours and the greatest response in the 500 mg group. Over 90 days, NAD^+^ concentrations increased in a time-and dose-related manner, with the 500 mg group showing an approximately threefold increase from baseline. The magnitude of whole-blood NAD^+^ augmentation was substantial; however, cross-trial comparisons with NMN or NR should be interpreted cautiously because of differences in compound chemistry, dose, assay methodology, biological matrix, study duration, and participant characteristics (Okabe et al., 2022) (Huang, 2022). Future studies should directly measure NMNH, NADH, NAD^+^, methylated metabolites, and related intermediates to better define the metabolic fate of NMNH-Ca in humans.

Albeit exploratory in nature, the geroscience-related endpoints of the present study suggested potential physiological relevance beyond NAD^+^ augmentation that require cautious interpretation. Estimated blood phenotypic age decreased most prominently in the 500 mg group, whereas an increase was observed in the placebo group. Because phenotypic age is a biomarker-derived estimate associated with morbidity and mortality risk in population studies, these findings should be interpreted as changes in systemic biomarker profile rather than direct evidence of biological age reversal (Z. Liu et al., 2018). The improvement in treadmill-based six-minute walk distance, particularly in the 500 mg group, was also notable, and the ≥50 m responder threshold was intentionally conservative relative to commonly reported minimal clinically important differences (Perera et al., 2006) (Myhre et al., 2024). Concordance between the treadmill-based 6MWT findings and selected SF-36 domains provides supportive, although not confirmatory, evidence that measured changes in functional performance may have been reflected in participants’ perceived physical health and vitality. In particular, improvements in the SF-36 physical functioning and energy/fatigue domains are directionally consistent with the observed increase in walking distance, suggesting possible alignment between objective functional testing and patient reported health status. However, these findings should be interpreted cautiously. Several SF-36 domains showed baseline imbalance across treatment groups, including domains related to bodily pain, social functioning, energy/fatigue, and general health, which may reflect chance imbalance due to modest group sizes, heterogeneity in baseline perceived health, or the inherently subjective nature of patient-reported outcomes. These baselines differences may have introduced residual confounding and limit the robustness of between-group interpretation. Accordingly, the SF-36 results should be regarded as exploratory and hypothesis-generating. Future trials should incorporate larger samples sizes, baseline stratification or baseline-adjusted analyses for patient-reported outcomes, standardized physical performance batteries, longer follow-up and mechanistic assessments to determine whether NAD^+^ augmentation produces reproducible improvements in both measured functional capacity and perceived health status.

Several limitations constrain interpretation and should be considered. First, this was a phase I study primarily designed to assess safety and tolerability, and exploratory endpoints were not powered to establish definitive clinical efficacy. Second, the 90-day intervention limits conclusions regarding durability, post-treatment persistence, and long-term safety. Third, the study population was relatively homogeneous, consisting entirely of Asian participants and predominantly male participants, which limits generalizability and precludes meaningful sex-specific inference. Fourth, whole-blood NAD^+^ may not reflect NAD^+^ dynamics in skeletal muscle, liver, brain, adipose tissue, or other aging-relevant organs. Finally, phenotypic age, six-minute walk distance, and SF-36 scores are susceptible to biological, behavioral, and measurement variability. Larger and more diverse trials incorporating tissue-relevant NAD^+^ metabolism, inflammatory and metabolic biomarkers, mitochondrial function, validated aging biomarkers, and comprehensive physical performance testing will be needed to determine whether the pharmacodynamic effects observed here translate into durable clinical benefit.

## 5. Conclusions

Oral NMNH-Ca supplementation was generally and well tolerated in healthy adults at doses up to 500 mg/day for 90 days, with no major short-term safety signal detected. NMNH-Ca produced rapid and sustained increases in whole-blood NAD^+^, with the largest pharmacodynamic response observed at 500 mg/day. Exploratory findings suggested favorable changes in blood phenotypic age, treadmill-based functional performance, and selected health-related quality of life domains; however, these outcomes are hypothesis-generating and require confirmation in larger, longer, and more diverse clinical trials with NAD^+^ metabolomics and comparator NAD^+^ precursor arms.

## Data Availability

All data produced in the present study are available upon reasonable request to the authors

## Acknowledgements

We thank iDD Research Solutions for their support in clinical trial coordination, monitoring, data management, regulatory compliance, quality control, biostatistical analysis, and biomarker assessment. We also thank all study participants and staff for their contributions.

## Author contributions

Conceptualization, Q.S., J.Y., E.J.; Data curation, Y.H.; Funding acquisition, Q.S., J.Y., E.J.; Investigation, J.L.; Methodology, J.L., Y.W., Y.L.; Supervision, R.K.; Visualization, J.L., Y.W., Y.L.; Writing—original draft preparation, J.L., Y.W., M.C.; Writing—review and editing, R.K., C.L..

All authors have read and agreed to the published version of the manuscript.

## Data Availability Statement

The data that support the findings of this study are available on request from the corresponding author. The data are not publicly available due to privacy or ethical restrictions.

