## Supplementary Document 1 for "Safety and Exploratory Efficacy of Reduced β-Nicotinamide Mononucleotide Calcium Salt (NMNH-Ca) in Healthy Middle-Aged and Older Adults: A Randomized, Double-Blind, Placebo-Controlled Trial"

**Table 1.** Summary of SF-36 scores

| Duration (Days) | Mean scores (± SD) | | | |
| --- | --- | --- | --- | --- |
|  | Placebo  (n = 21) | NMNH-Ca  125 mg  (n = 18) | NMNH-Ca  250 mg  (n = 20) | NMNH-Ca  500 mg  (n = 21) |
| Physical function | | | | |
| Baseline | 63.95 ± 4.81 | 63.22 ± 4.31 | 63.90 ± 6.38 | 61.90 ± 5.06 |
| Day 45 | 66.00 ± 7.75 | 70.94 ± 7.8*** | 73.65 ± 6.38*** | 73.57 ± 9.37*** |
| Day 90 | 66.19 ± 9.40 | 67.67 ± 17.89 | 77.50 ± 6.04*** | 79.29 ± 7.96*** |
| Body pain | | | | |
| Baseline | 55.26 ± 10.95 | 66.03 ± 5.14 | 66.88 ± 10.79 | 63.31 ± 6.00 |
| Day 45 | 56.98 ± 12.13* | 69.39 ± 3.57*** | 74.75 ± 11.68*** | 70.71 ± 7.79*** |
| Day 90 | 57.10 ± 10.57 | 67.00 ± 17.44 | 79.35 ± 12.76*** | 74.62 ± 8.75*** |
| Role limitations due to physical health problems | | | | |
| Baseline | 60.81 ± 6.61 | 59.72 ± 4.01 | 63.65 ± 7.23 | 62.05 ± 6.09 |
| Day 45 | 59.48 ± 9.24 | 60.28 ± 4.16 | 74.75 ± 5.73*** | 71.57 ± 4.73*** |
| Day 90 | 62.10 ± 7.50* | 65.67 ± 4.20*** | 77.10 ± 5.41*** | 75.38 ± 6.17*** |
| Role limitations due to emotional problems | | | | |
| Baseline | 61.11 ± 7.04 | 66.45 ± 3.86 | 65.02 ± 3.24 | 63.84 ± 4.71 |
| Day 45 | 60.91 ± 8.26 | 70.15 ± 2.64*** | 75.18 ± 7.52*** | 78.50 ± 7.38*** |
| Day 90 | 57.73 ± 19.11 | 72.09 ± 3.04*** | 77.60 ± 7.55*** | 82.95 ± 6.73*** |
| Emotional well-being | | | | |
| Baseline | 58.95 ± 10.15 | 66.39 ± 9.31 | 64.95 ± 7.46 | 65.52 ± 9.72 |
| Day 45 | 60.00 ± 11.44 | 70.11 ± 8.13*** | 77.55 ± 10.46*** | 78.52 ± 11.89*** |
| Day 90 | 63.48 ± 11.40** | 75.06 ± 6.10*** | 81.70 ± 8.23*** | 85.55 ± 10.96*** |
| Social function | | | | |
| Baseline | 56.29 ± 8.53 | 68.64 ± 8.62 | 62.23 ± 12.20 | 67.98 ± 12.28 |
| Day 45 | 58.49 ± 7.48** | 71.53 ± 7.06*** | 72.20 ± 11.56*** | 80.52 ± 12.61*** |
| Day 90 | 59.45 ± 10.83 | 74.47 ± 5.91*** | 76.00 ± 12.12*** | 85.14 ± 10.19*** |
| Energy/fatigue | | | | |
| Baseline | 49.10 ± 10.37 | 62.56 ± 10.82 | 58.45 ± 13.55 | 63.81 ± 11.39 |
| Day 45 | 52.95 ± 12.73* | 67.61 ± 7.69*** | 73.00 ± 14.43*** | 78.43 ± 13.41*** |
| Day 90 | 52.86 ± 15.50 | 71.56 ± 6.67*** | 78.25 ± 13.43*** | 86.24 ± 12.66*** |
| General health | | | | |
| Baseline | 57.67 ± 7.34 | 66.61 ± 8.56 | 69.90 ± 9.16 | 70.33 ± 8.48 |
| Day 45 | 61.86 ± 7.95*** | 72.53 ± 8.81*** | 76.10 ± 8.43*** | 78.95 ± 6.27** |
| Day 90 | 63.57 ± 10.64*** | 75.94 ± 7.20*** | 79.35 ± 9.99*** | 84.95 ± 7.87** |

Values are presented as mean ± SD. Within-group comparisons between baseline and each post-baseline time point were performed using paired *t*-tests. **p* < 0.05, ***p* < 0.01, and ****p* < 0.001 versus baseline.

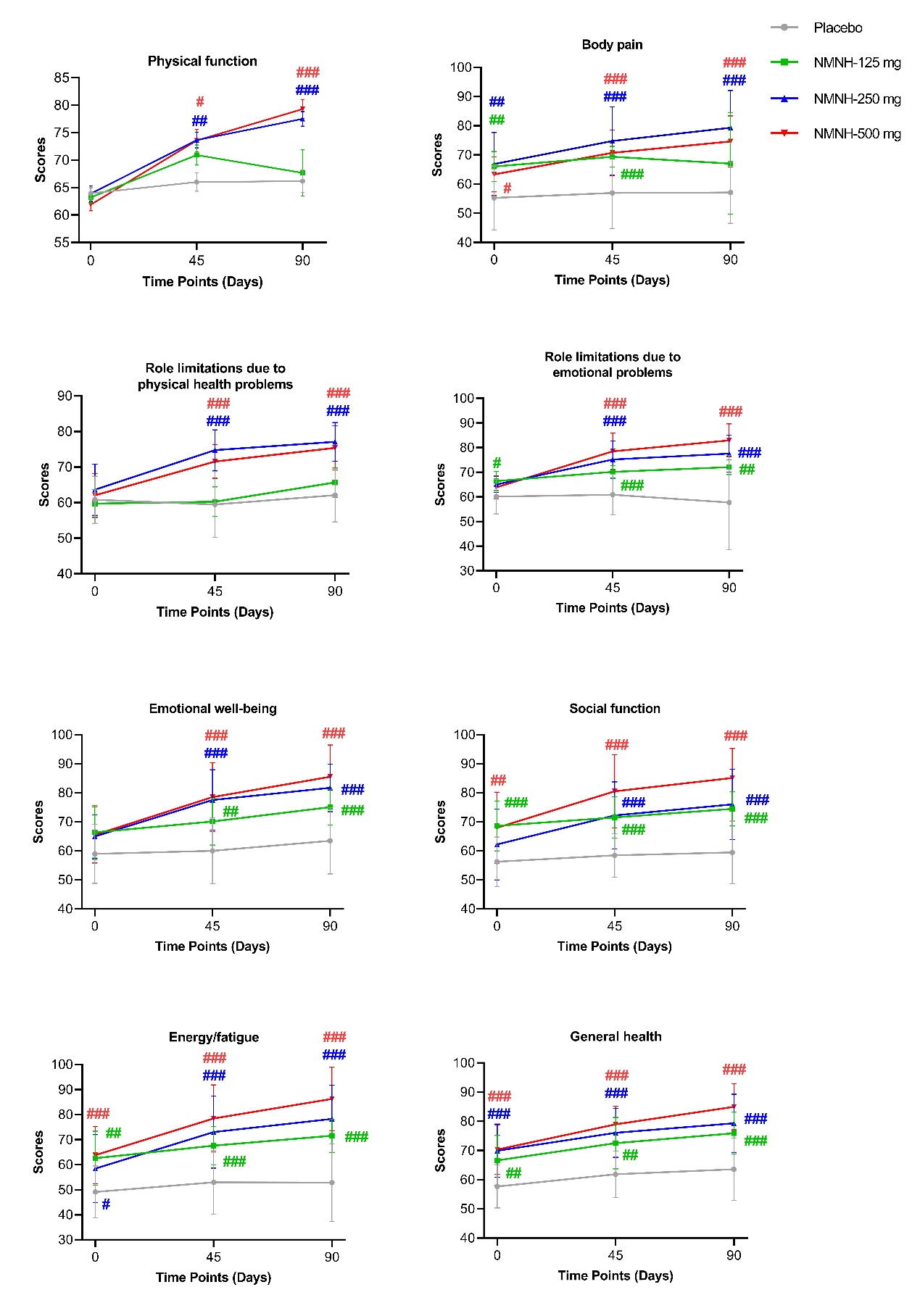

**Figure 1**. Effects of NMNH-Ca supplementation on SF-36 domain scores over 90 days.

SF-36 domain scores were assessed at baseline, Day 45, and Day 90 in participants receiving placebo or NMNH-Ca at doses of 125, 250, or 500 mg. The eight domains included physical function, body pain, role limitations due to physical health problems, role limitations due to emotional problems, emotional well-being, social function, energy/fatigue, and general health. The placebo group is shown in gray, the NMNH-Ca 125 mg group in green, the NMNH-Ca 250 mg group in blue, and the NMNH-Ca 500 mg group in red. Higher scores indicate better health-related quality of life. Data are presented as mean ± SD. Between-group comparisons between each NMNH-Ca group and the placebo group over time were performed using a mixed model for repeated measures. #*p* < 0.05, ##*p* < 0.01, and ###*p* < 0.001 versus placebo.
