## Supplementary Document 2 for "Safety and Exploratory Efficacy of Reduced β-Nicotinamide Mononucleotide Calcium Salt (NMNH-Ca) in Healthy Middle-Aged and Older Adults: A Randomized, Double-Blind, Placebo-Controlled Trial"

**Supplementary document 1**

Detailed description of the definitions listed in Table 6.

**Availability of treatment:**

Adverse events (AEs) requiring therapy must be treated by recognized standards of medical care to protect the health and well-being of the Subject. Appropriate resuscitation equipment and medicines must be available to ensure the best possible treatment of an emergency situation.

**The outcome of AEs will be rated as:**

- complete recovery
- incomplete recovery
- unknown
- Death

**Causality:**

The following five-point scale will be used for rating the causal relationship of the AEs to the investigational product:

| Unrelated: | Clearly and incontrovertibly due only to extraneous causes, and does not meet criteria listed under unlikely, possible or probable. |
| --- | --- |
| Unlikely: | Does not follow a reasonable temporal sequence from administration. May have been produced by the Subject's clinical state or by environmental factors or other therapies administered. |
| Possible: | Follows a reasonable temporal sequence from administration. May have been produced by the Subject's clinical state or by environmental factors or other therapies administered. |
| Probable: | Clear-cut temporal association with improvement on cessation of test drug or reduction in dose. |
| Certain: | Clear-cut temporal association with improvement on cessation of test drug or reduction in dose. Reappears upon re-challenge. Follows a known pattern of response to test drug. |
